# Prevalence of and risk factors for burnout and occupational stress among couriers: A systematic review

**DOI:** 10.1101/2021.11.09.21266103

**Authors:** Hua Wei, Shugang Li, Sheng Li, Thomas O’Toole, Mengke Yu, Christopher J. Armitage, Tarani Chandola, Pauline Whelan, Yan Xu, Martie van Tongeren

**Affiliations:** Division of Population Health, Health Services Research & Primary Care, School of Health Sciences, University of Manchester, Manchester, UK; Department of Maternal and Children Health, School of Public Health, Capital Medical University, Beijing, China; Faculty of Psychology, Beijing Normal University, Beijing, China; Department of Public Health, School of Medicine, Shihezi University, Shihezi, China; Division of Psychology & Mental Health, School of Health Sciences, University of Manchester, Manchester, UK; Department of Sociology, The University of Hong Kong, Hong Kong SAR China; Division of Informatics, Imaging & Data Sciences, School of Health Sciences, University of Manchester, Manchester, UK

**Author notes:** **Correspondence:** Hua Wei, The University of Manchester, Ellen Wilkinson Building, Oxford Road, Manchester, M13 9PL.

**Keywords:** burnout, occupational stress, couriers, gig economy, China

## Abstract

**Objective:** To estimate the prevalence of burnout and occupational stress (OS) among couriers and to identify the main risk factors.

**Method:** We followed PRISMA guidelines to search studies published in English and Chinese databases before February 2022.

**Results:** The search yielded 15 (7 English and 8 Chinese) papers, 12 of which studied Chinese couriers, and three studied French, Israeli and Malaysian couriers. Twelve studies reported OS using various validated measurement tools, and six reported burnout using adapted versions of Maslach Burnout Inventory. Only four included studies suggested cut-off points to define the condition, hence we used the midpoint of the scales to estimate the prevalence (the proportion of the cases that were above the midpoint). The estimated prevalence of burnout (or a dimension of burnout) among couriers ranged from 20% to 73% (median=33%). The prevalence of OS (or a dimension of OS) ranged from 7% to 90% (median=40%). Twelve studies reported risk factors for burnout or OS; the main ones were physical demands, customer behaviour and a range of working and employment conditions, such as employment precarity and financial insecurity. Job resources (i.e. social support and decision latitude) and organizational support had mitigating effects.

**Conclusions:** Burnout and OS are relatively high among couriers. Interventions to prevent or reduce burnout in this occupation are currently limited. The use of platform technology to shift risks, intensify work and tighten managerial controls could be a potential work-related stressor for app-based couriers but remains a knowledge gap.

## BACKGROUND

The rise of the gig economy and the post-COVID preference for socially distanced working and living have continued to boost global e-commerce sales and the delivery sector. EMarketer, the digital market research company, estimated that retail e-commerce sales worldwide experienced a 25.7% surge in 2020, to $4.213 trillion, and climbed a further 16.8% in 2021, to $4.921 trillion.^1^ This dramatic growth has led to increased demands and pressure for the logistics sector and delivery workers. Courier jobs are often characterised as high demand, fast-paced, low control with lack of support, indicating that work stress and job burnout could be high within this occupation (Bakker & Demerouti, 2017; Kristensen et al., 2005; Shoman et al., 2021). However, there is a paucity of knowledge about whether and how this population are stressed at work. The main aim of this review is to estimate the prevalence of couriers’ burnout and occupational stress (OS). We will also qualitatively synthesise the main risk and mitigating factors that could inform the development of preventative measures in future research.

The burnout and OS issues among couriers have come to our attention not just because of the increased demand for home deliveries during the COVID-19 pandemic, but also because of the rapid expansion of e-commerce in developing countries (Tediosi et al., 2020). E-commerce markets in China, India, Brazil, Russia, and Argentina are all projected to achieve two digits growth in 2021, according to eMarketer.^2^ Furthermore, parcel and food delivery is an essential part of the gig economy, where workers are categorised as self-employed and the platforms do not provide the traditional employer responsibilities, such as health, safety and other employee benefits. While this mode of working has the benefit of flexibility, analysis of large scale survey data in North America has shed light on the relationship between gig workers’ socio-economic status and work stress, that is the dependency on platforms (ie as the main job and only source of income) and financial strain exacerbate the mental health penalties associated with platform work (Glavin & Schieman, 2021).

Surging demand, relatively underdeveloped labour protection, employment insecurity and economic precarity could all add to the stressful work environment for the couriers (Karasek et al., 1998). China is the global leader of e-commerce and the fastest growing region. Its outsized role means that the Asia-Pacific region accounted for 60.8% of retail ecommerce sales worldwide in 2021, while North America had a 20.3% share, and Western Europe 12.6%.^3^ The phenomenon has subsequently triggered a large amount of research published in Chinese. Research about overwork among Chinese takeaway couriers found that 41.7% respondents (n=1,114) were at high risk of overworking, 35.9% were at very high risk of overworking and only 7.9% were considered risk free (Lin & Li, 2021). A recent road safety study from China revealed that 76.5% of interviewed couriers (n=480) had been involved in a traffic crash at least once, while the average length of staying in this occupation was only about 1.5 years (Z. Wang et al., 2021). These results are alarming and are suggestive of chronic and excessive work stress in this population. The Chinese language literature may therefore be a key area to search for relevant research on burnout in couriers and will go some way to extending the reach of previous systematic reviews of burnout, which to date have focused on English language databases, effectively ignoring vast sections of the global economy.

It is important to describe the prevalence of burnout and OS in this occupational group, prior to the development of interventions to prevent such detrimental effects on workers’ mental and physical health in the long term. This review aims answer the questions by reviewing evidence from the Chinese and English language literatures. Our review questions are:

Q1 What is the prevalence of job burnout and occupational stress among couriers?
Q2 What are the main exacerbating and ameliorating factors of job burnout and occupational stress among couriers?

## METHOD

### Search strategy

The searches were completed in February 2022 to catch the latest publications.

We searched the following electronic databases for English literature:

- MEDLINE, EMBASE, PsycINFO and the Cochrane Library (from their respective inception dates to the current date) using OVID platform.
- Web of Science (Core Collection)
- Pre-print database: Open Science Framework
- Grey Literature database: Google Scholar

We also searched the following Chinese electronic databases for Chinese literature:

- CNKI (http://new.oversea.cnki.net/index/).
- WANFANG data
- Sino Med
- VIP (http://www.cqvip.com/)

For search terms on OVID, we started with Exp Burnout to catch as much as possible relevant terms, in addition, we used (burnout OR stress OR job strain OR fatigue OR exhaustion OR tired OR tiredness OR tiring OR weariness OR worn out) to include possibly related terms in social and health sciences. For the occupation, we used (courier* or deliverym*n or delivery worker* or delivery driver* or takeaway delivery) to catch all relevant population in focus. At the end, we used (work OR occupational OR job) to limit results to studies that are work-related. A full search strategy on all databases is available as supplementary material.

We also hand searched the reference lists of all the included studies, and of some excluded studies. We have reviewed peer-reviewed papers and postgraduate theses that are published in English or Chinese, with the full text available. No restriction on publication dates were applied.

### Inclusion/Exclusion criteria

Reviews, RCT, CRT, observational and qualitative studies are all eligible.

Inclusion criteria: Studies reported level of burnout or OS, or relevant risk factors in a population of couriers/delivery workers.

Exclusion criteria: Studies not reporting work-related mental ill health or not in a sample of delivery workers or couriers; Conference abstracts; Commentaries; Studies not in English or Chinese language.

Couriers including parcel/goods couriers and takeaway food couriers who conduct the “last mile” deliveries will be the focus of the review. Hence, studies about postmen, public transport drivers and long-haul truck or lorry drivers were excluded.

### Data extraction

We followed a review process adapted from the Preferred Reporting Items for Systematic Reviews and Meta-Analyses (Stevens et al., 2018) (see Figure 1 PRISMA diagram below). A review protocol was pre-published on PROSPERO (ID: CRD42021247644).

**Figure 1.**
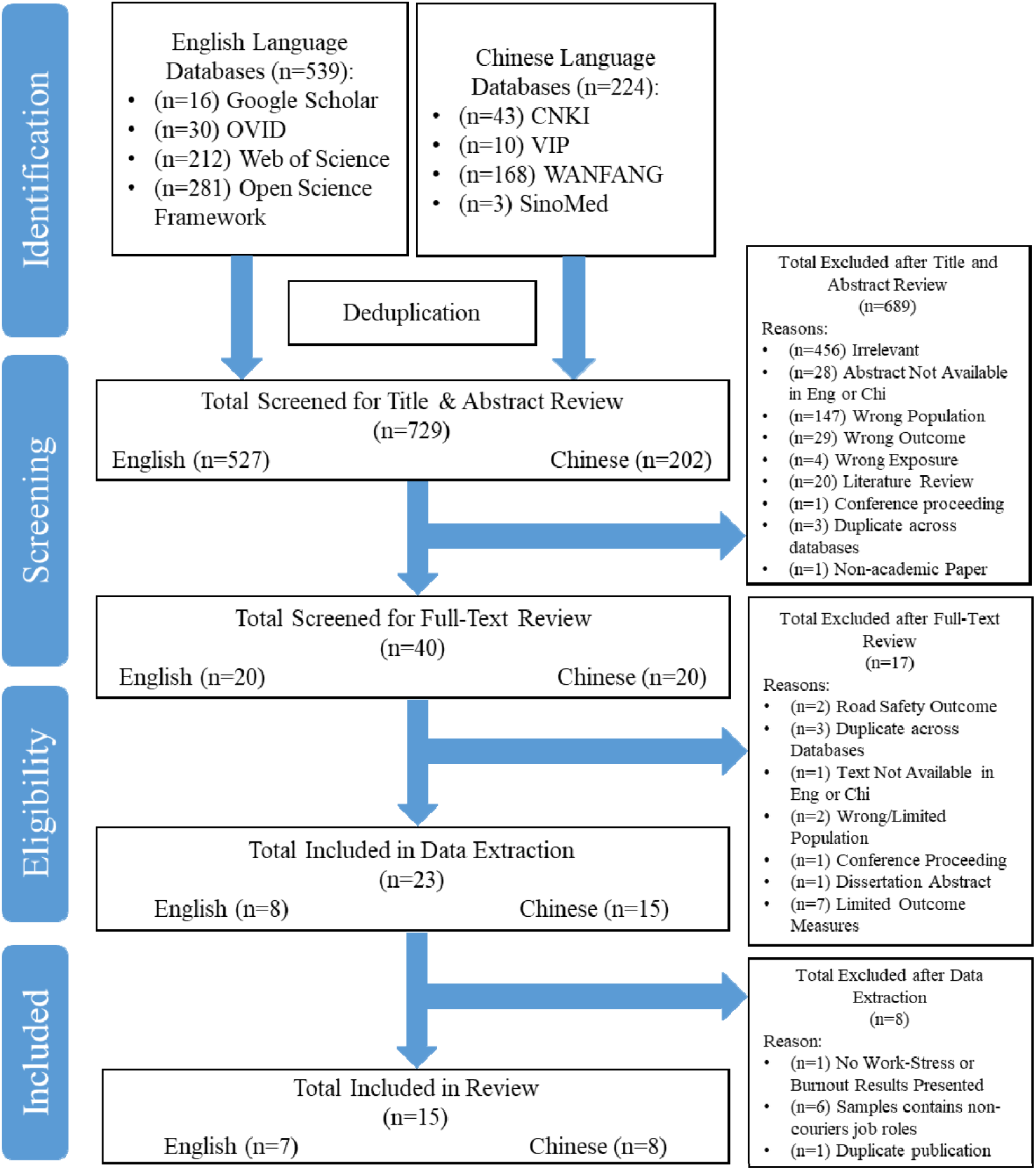
PRISMA diagram

Five reviewers (HW, TO, SGL, SL, MKY) formed two teams to carry out systematic reviews in Chinese and English. Author SGL, SL and MKY were responsible for Chinese review and HW and TO English. Both teams went through the same steps, including searching the databases, screening the results, data extraction and quality assessment. The protocol was reviewed by and discussed among all the co-authors, with feedback actively taken into account to ensure quality of the process. HW and SGL cross checked both languages at each step to maintain consistent standard. When both reviews were completed, HW translated data extracted from the Chinese review into English. The abstracts of all included Chinese studies were also translated into English by author HW to enable discussions among all co-authors.

We used standardised forms for title/abstract screening, full-text review and data extraction. The forms were developed in English by all five reviewers. Each team conducted pilot exercises with the same 10 articles to calibrate. Any conflicts were discussed and if disagreement remained, we would refer to the review team for further discussion until consensus was reached. Two reviewers of each team then completed reviewing or extracting the rest of the literature independently with conflict resolution through discussion and referral to a third reviewer if necessary.

The same process was followed for risk of bias assessments. A version of the Newcastle Ottawa Scale (NOS) that was adapted for cross-sectional surveys and cohort longitudinal studies were used for quality assessment (Herzog et al., 2013). The Critical Appraisal Skills Programme (CASP) checklists were used for qualitative studies.

### Data synthesis and analysis

We planned to perform meta-analysis of the prevalence. However, due to the heterogeneity in burnout and OS ascertainment methods, definitions (or lack of), scoring scales, bias from low quality studies and limited number of eligible studies, the pooled quantitative results were judged to be unreliable. Qualitative synthesis was deemed the most appropriate method and the included studies were summarized descriptively with outcomes of interest displayed in tables.

### Cut-off points and definition of prevalence

None of the included studies on Burnout provided cut-off points to determine the outcome. The absence of or lack of agreement on cut-off scores appears to be a common problem in burnout research. Rotenstein et al. (2018) reviewed 182 studies on physicians’ burnout and reported that about one third of the studies did not specify cut-off scores and the specified cut-off scores in the rest of the studies varied significantly. The most commonly used cut-off scores were similar to the original MBI manual suggestion: EE≥27 (total 54) and DP≥10 (total 30) and PA≤33 (total 48), but were only used by 11% of the 182 studies (Maslach et al., 1997; Rotenstein et al., 2018). In our searches, one study that was excluded at the stage of data extraction (due to a mixed courier and non-courier sample) specified the midpoint of the total scores as the a cut-off (Zhao, 2020). Four studies on OS specified cut-off points. Wang et al. (2015) used midpoint of the scoring scale 3 (Likert 1-5). Jiang (2017) used Standard T scores to determine moderate and high of three subscales. Lin and Li (2021) defined that when the ratio of work demand subscale to job autonomy subscale >1, OS was high. Egozi et al. (2021) used a Likert 1-5 scoring scale and determined that mean score >4 indicated high stress.

Since only four included studies suggested cut-off points to define the condition, we used the midpoint of the scales to estimate the prevalence. The prevalence is defined as the estimated proportion of cases scored above midpoint. Standard Z score is calculated using equation z = (x – μ) / σ, where x refers to midpoint, μ refers to mean and σ refers to standard deviation (SD). We then used Z score of the normal distribution principle to deduce the area corresponding to the midpoint (that is, the proportion of cases scored below midpoint) and the prevalence was calculated as “1-area value” (the proportion of cases scored above midpoint).

## RESULTS

### Search results

Databases searches returned 763 results and 34 were removed after deduplication. In total 729 papers entered title/abstract screening, 527 of which were in English and 202 in Chinese. Following title and abstract screening, 40 papers were included for full-text review, after which 17 were further excluded, leaving 23 papers for data extraction. During data extraction, a further 8 papers were excluded, mostly because the sample contained non-courier job roles. At the end, 15 papers were included for reporting and quality assessment. More detailed reasons of exclusion at each stage are illustrated in Figure 1.

### Study characteristics

Fifteen studies (thirteen cross-sectional, one longitudinal and one qualitative) were included, reporting findings from mainland China, Taiwan, France, Israel and Malaysia. Seven papers were published in English and eight in Chinese. The number of participants ranged from 32 to 2887, with an average of 832 per study. Samples were primarily male (∼16% female on average), with the most frequent age band reported being ∼20-40. One study did not report age or sex/gender statistics. Six papers studied burnout or a dimension of burnout, and twelve papers studied OS or a dimension of OS, with three papers researched both. Twelve papers reported risk or mitigating factors of burnout or OS. Table 1 describes the characteristics of studies included in this review.

**Table 1.** Study characteristics

| Reference | Country, language | Study type | Job type | Sample size | Age (mean/band)** | % Female | Outcome of interest |
| --- | --- | --- | --- | --- | --- | --- | --- |
| Cheng & Chen (2020) | China, E | Cross-sectional | Parcel | 220 | 30.9% (26-30) | 14% | EE, risk factors |
| Yoon et al (2021) | Malaysia, E | Cross-sectional | Parcel | 350 | 50% (21-40) | 10% | Burnout, risk factors |
| Apouey et al (2020) | France, E | Cross-sectional | Mixed | 52 | Driver 50% (50+),<br>Biker 67.7% (18-24) | Driver 0%,<br>Biker 6.7% | OS, risk factors |
| Lin et al (2011) | Taiwan, E | Cross-sectional | Parcel | 32 | 38.0 ± 8.8 | 0% | OS, risk factors |
| Xu & Zhu (2020) | China, C | Cross-sectional | Takeaway | 160 | 75% (20-40) | 25% | OS, EE, DP, LSA, risk factors |
| Wang et al (2015) | China, C | Cross-sectional | Parcel | 686 | NR | NR | OS, Burnout, risk factors |
| Jiang (2017) | China, C | Cross-sectional | Mixed | 322 | 33.5% (30-40) | 23% | ORQ, PSQ, PRQ |
| Xiao (2019) | China, C | Cross-sectional | Mixed | 1425 | 92.2% <42 | 8% | Burnout, risk factors |
| Li et al (2021) | China, C | Longitudinal cohort | Takeaway | 130 | 27.6±5.1 | 9% | PR, PsyR, BR, WP, risk factors |
| Lian & Zhou (2019) | China, C | Qualitative | Takeaway | 1692 | 16-35 | NR | OS, risk factors |
| Yang & Mo (2021) | China, E | Cross-sectional | Mixed | 348 | 51.9% (18-33) | 18% | OS |
| Lin & Li (2021) | China, C | Cross-sectional | Takeaway | 1114 | 80.61% (20-34 ) | 3% | OS |
| Egozi et al (2021) | Israel, E | Cross-sectional | Mixed | 237 | 25.2 | 33% | OS, risk factors |
| Xie, Tian et al (2021)* | China, E | Cross-sectional | Parcel | 2831 | 40.6% (31–40) | 26% | OS, risk factors |
| Xie, Jiao et al (2021) | China, C | Cross-sectional | Parcel | 2887 | 34.63±8.63 | 26% | OS, Burnout, risk factors |

Abbreviations: NR = Not Reported, E: English, C: Chinese, EE: Emotional Exhaustion, DP: Depersonalisation, LSA: Low sense of accomplishment, OS: Occupational Stress, ORQ: Occupational Role Questionnaire, PSQ: Personal Strain Questionnaire, PRQ: Personal Resources Questionnaire, PR: Physical response, PsyR: Psychological response, BR: Behavioural response, WP: Work performance
\*Sample partially overlaps with Xie, Jiao et al 2021. \*\*When mean age was not reported, the biggest age band was used

### Quality assessment

Table 2 shows the results of various quality assessments carried out on the included studies. Across the studies, quality ranged from 3/10 to 7/10. Most systematic reviews consider 6 or 7 and above as high quality (Daniels et al., 2021; Farsad-Naeimi et al., 2020; J. Wang et al., 2021). In Table 2, we found six papers (bold in the table) that can be described as good quality whilst the others were of low or medium quality.

**Table 2.** Quality assessment of the included studies

| Reference | Quality assessment tool | Score |
| --- | --- | --- |
| Cheng et al, 2020 | NOS Cross-Sectional | 4/10 |
| Yoon et al, 2021 | NOS Cross-Sectional | 5/10 |
| Apouey et al, 2020 | NOS Cross-Sectional | 3/10 |
| Lin et al, 2011 | NOS Cross-Sectional | 5/10 |
| Xu & Zhu, 2020 | NOS Cross-Sectional | 4/10 |
| Wang et al, 2015 | NOS Cross-Sectional | 5/10 |
| **Jiang, 2017** | **NOS Cross-Sectional** | **7/10** |
| **Xiao, 2019** | **NOS Cross-Sectional** | **6/10** |
| Li et al, 2021 | NOS Longitudinal | 4/9 |
| **Lian & Zhou, 2019** | **CASP** | **7/10** |
| Yang & Mo, 2021 | NOS Cross-Sectional | 4/10 |
| **Lin & Li (2021)** | **NOS Cross-Sectional** | **6/10** |
| Egozi et al (2021) | NOS Cross-Sectional | 5/10 |
| **Xie, Tian et al (2021)\*** | **NOS Cross-Sectional** | **7/10** |
| **Xie, Jiao et al (2021)** | **NOS Cross-Sectional** | **6/10** |

Abbreviations: NOS Cross-Sectional = Newcastle Ottawa Scale; NOS Longitudinal = Newcastle Ottawa Scale for Cohort longitudinal studies; CASP = Critical Appraisal Skills Programme
\*Sample partially overlaps with Xie, Jiao et al 2021.

### Measurements used to assess burnout and OS

All six included studies on burnout used a version of Maslach Burnout Inventory (MBI), or a subscale of MBI, or when the scale was unspecified, the dimensions appeared to be consistent with MBI. However, the measurement versions and scoring scales varied significantly (see table 3.1 and 3.2 for details). Twelve studies were included that researched OS among couriers. Ten of them reported OS scores using validated OS measurements. The only two papers that used the same scale were Xie, Tian et al (2021) and Xie, Jiao et al. (2021). These two papers both used the Chinese work stress scale (CWSS) but their samples potentially overlapped with each other because the two studies appeared to be based on the same dataset with different inclusion criteria.

### Prevalence of burnout and OS among couriers

The estimated prevalence of burnout (or a dimension of burnout) among couriers ranged from 20% to 73% (median=33%). The prevalence of OS (or a dimension of OS) ranged from 7% to 90% (median=40%). Most papers did not explicitly state whether their data was normally distributed. In which case, we assumed normality when mean and SD were used to describe the data. Table 3.1 and 3.2 present the prevalence of burnout and OS using the midpoint as cut-off. Two studies, Cheng and Chen (2020) and Wang et al. (2015) did not report burnout scores and were not included in table 3.1. Three papers were not included in table 3.2 for analysis, because Apouey et al. (2020) and Lian and Zhou (2019) did not report mean and SD of their results and Lin et al. (2010) had a sample size (n=32) that was too small to meet the normal distribution assumption.

**Table 3.1.** Measurements and prevalence of burnout or dimensions of burnout

| Reference | Outcome | Sample size | Measurement tools | Scoring scale | Cut-off | Midpoint | Mean | SD | Normality | Z score | Area to the left | Prevalence* |
| --- | --- | --- | --- | --- | --- | --- | --- | --- | --- | --- | --- | --- |
| Xu & Zhu, 2020 | EE | 160 | 12-item Chinese aMBI, 4-item EE | Likert 1-5 | NR | 12 | 10.775 | 2.5648 | Assumed | 0.48 | 0.68439 | 32% |
|  | DP | 160 | 12-item Chinese aMBI, 4-item DP | Likert 1-5 | NR | 12 | 10.5 | 2.3871 | Assumed | 0.63 | 0.73565 | 26% |
|  | LSA | 160 | 12-item Chinese aMBI, 4-item LSA | Likert 1-5 | NR | 12 | 10.975 | 2.4622 | Assumed | 0.42 | 0.66276 | 34% |
| Xiao, 2019 | Burnout | 1425 | 15-item Chinese aMBI-GS -Li | Likert 0-7 | NR | 3.5 | 4.02 | 0.85 | Assumed | -0.61 | 0.27093 | 73% |
| Xie, Jiao et al 2021 | EE | 2887 | 5-item MBI-GS, EE dimension | Likert 0- 6 | NR | 15 | 13.42 | 5.53 | Assumed | 0.29 | 0.61409 | 39% |
| Yoon et al, 2021 | Burnout | 350 | 16-item aMBI-GS, Bahasa Malaysia version | Four-point 1-4 | NR | 3 | 2.45 | 0.651 | Univariate normality | 0.84 | 0.79955 | 20% |

**Table 3.2.**
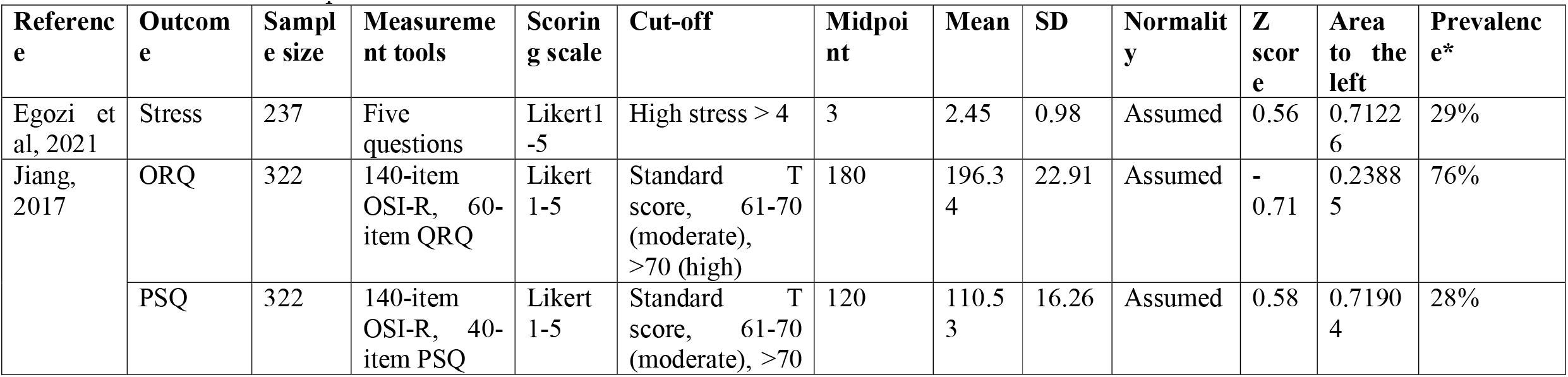
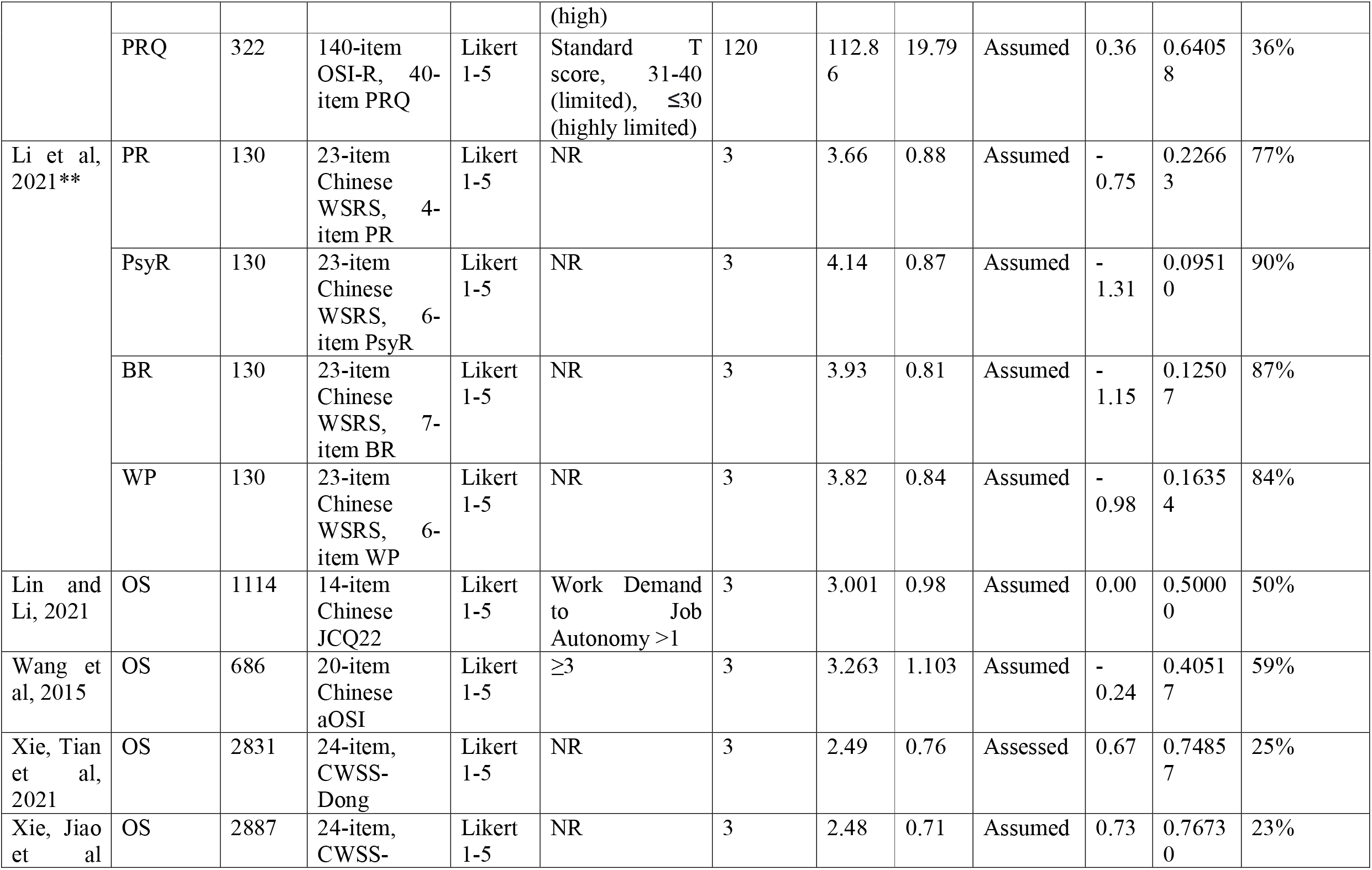
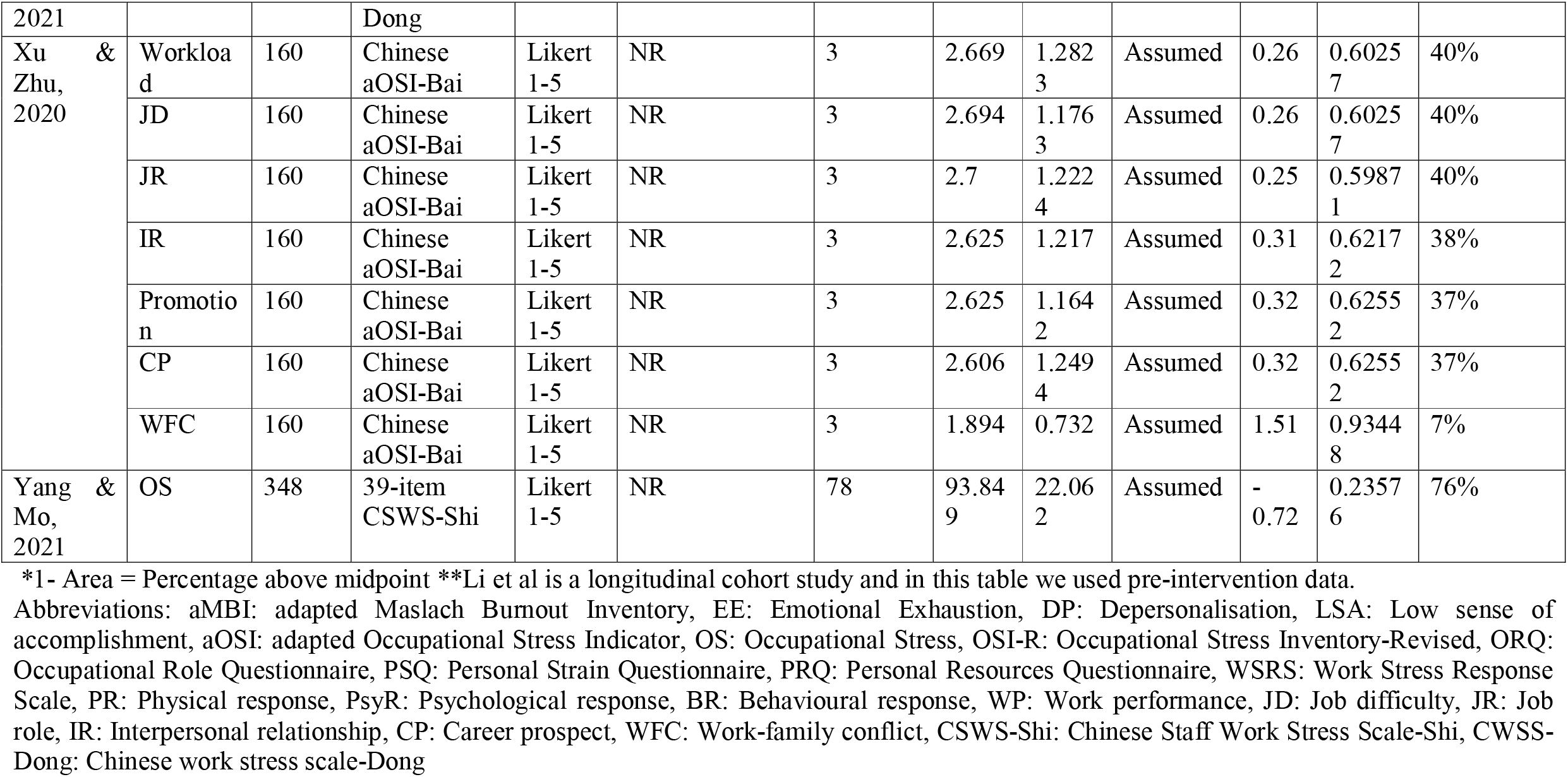
Measurements and prevalence of OS or dimensions of OS

Figure 2.1 and 2.2 show the forest plots and pooled prevalence of overall burnout and OS. When the studies provided Mean and SD of the dimensions rather than that of the overall scale, the prevalence of dimensions were estimated (using the number of cases above midpoint) and then pooled to estimate the overall prevalence. As explained in the Measurement section, due to the high heterogeneity and the limited number of eligible studies, the results were judged to be unreliable.

**Figure 1.**
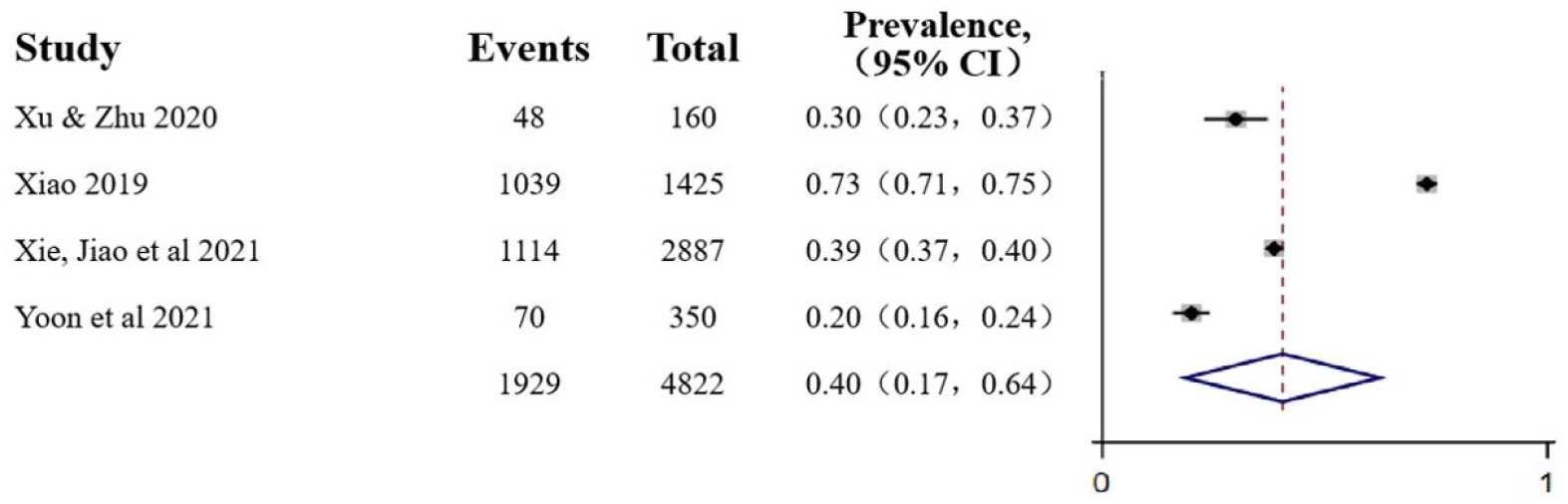
Pooled results of prevalence of overall burnout

**Figure 2.**
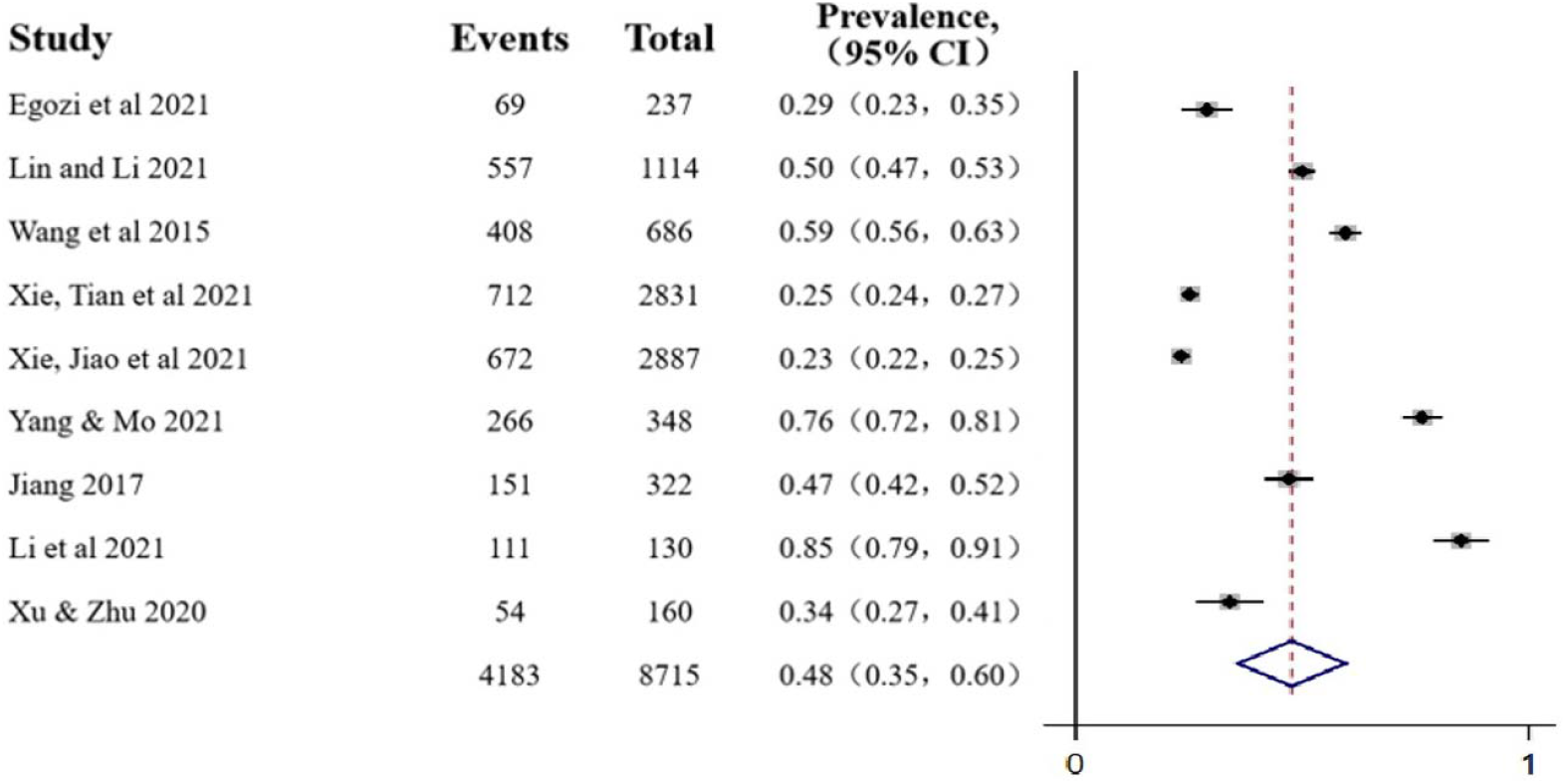
Pooled results of prevalence of overall OS

### Main factors of burnout and OS

All included studies reported one or more risk or mitigating factors for job burnout among couriers (see details in Table 4), with five of them reported in both categories. The relationships between the factors and burnout were established using statistical methods such as Correlation Coefficient (CC), structural equation modelling (SEM), or standardized coefficient (SC). Customer behaviour, physical job demands and OS are found to have negative impact on job burnout. Psychological empowerment, job resources (including its social support and decision latitude dimensions), and perceived organizational support are found to have mitigating effects against burnout. Age, education, years of working, marital status, income are among the socio-demographic factors that differentiating burnout experiences. Table 4.1 summaries the relevant factors.

**Table 4.1.** Risk and mitigating factors for burnout

| Category | Factors | Reference | Analysis methods | Results (rounded to two decimals) |
| --- | --- | --- | --- | --- |
| Exacerbating factors | Customer behaviour | Cheng & Chen | SEM | Improper customer interaction has a significant positive effect on EE ($\beta=0.61$ , T-value =9.38) |
| | Physical JD | Yoon et al | CC<br>CB-SEM | Job demand is correlated with job burnout $r=0.53, p<0.01$;<br>The result of the structural paths shows that higher job demands have increased burnout ($\beta = 0.52, p < 0.00$). However, only one of the job demands dimensions - physical demands - has created a positive effect on burnout ($\beta = 0.50, p<0.00$). |
|  | OS | Xu & Zhu | NR | Association between work stress and job burnout among couriers were not significant (Effect not reported) |
| | | Wang et al | Pearson CC | OS is significantly correlated with job burnout ($r=0.68, p<0.01$), and its three dimensions (EE 0.93, CD 0.85, LPE 0.90, $p<0.01$) |
| | | Xie, Jiao et al | Pearson CC | Work Stress is significantly positively correlated with EE ($r=0.429, P<0.01$) |
| Ameliorating factors | PE | Xiao | CC | PE is negatively correlated with burnout $r=-0.39, p<0.01$ |
| | JR | Yoon et al | CB-SEM, SPC | Job resources moderate the effect of job demands on burnout significantly ( $\beta=-0.30$ , p<0.000) |
| | JR -- SoS | Yoon et al | CB-SEM, SPC | SoS moderates the effect of physical and psychological job demands on burnout ($\beta=-0.27$, $p<0.000$, $\beta=-0.33$, $p<0.000$) |
| | JR -- DL | Yoon et al | CB-SEM, SPC | DL moderate the effect of physical and psychological job demands on burnout ($\beta=-0.26$, $p<0.000$, $\beta=-0.28$, $p<0.000$) |
| | POS | Cheng & chen | SEM | POS has a negative regulating effect between improper customer interaction and EE: high POS group $\beta = 0.12$, compared to low POS group $\beta = 0.48$, $p<0.001$ |
| | | Wang et al | Pearson CC | OS is significantly negatively correlated with job burnout (Pearson CC=-0.518, $p=0.01$). Taking work stress factors as independent variables, burnout dimensions as dependent variables, OS significantly moderate the effect between work stressors and burnout dimensions. |
| Socio-demographic factors | Age | Xiao, 2019 | T-test/ANOVA | MBI mean score: 26-33 vs 18-25 age group, $4.02\pm0.85$ vs $3.48\pm1.12$, $p<0.05$; |
| | | Xie, Jiao et al | T-test/ANOVA | EE: $\leq 30$ $13.58\pm5.12$ vs $31-40$ $13.62\pm5.59$ vs $>40$ $12.76\pm6.04$ ($p<0.01$) |
| | Education level | Xie, Jiao et al | T-test/ANOVA | EE: Lower middle school or below $12.39\pm5.55$ vs High school $13.72\pm5.76$ vs College $13.55\pm5.42$ vs Degree or above $14.41\pm4.66$ ($p<0.01$) |
| | Years of working | Xie, Jiao et al | T-test/ANOVA | EE: $\leq 4$ $13.18\pm5.38$ vs $5-10$ $13.54\pm5.75$ vs $>11$ $14.12\pm5.39$ ($p<0.05$) |
| | Marital status | Xiao, 2019 | T-test/ANOVA | MBI mean score: Married vs unmarried, $4.13\pm0.79$ vs $3.77\pm0.93$, $p<0.05$ |
| | Income level | Xiao, 2019 | T-test/ANOVA | MBI mean score: Monthly income $>5000$ vs $<3000$ group, $4.23\pm0.83$ vs $3.22\pm1.15$, $p<0.05$; |

Abbreviations: NR: Not reported, EE: Emotional Exhaustion, CD: Cynicism/Depersonalisation, LSA: Low sense of accomplishment, LPE: Low professional efficacy, OS: Occupational stress, JD: Job demands, JR: Job resources, SoS: Social Support, DL: Decision Latitude, PE: Psychological Empowerment, POS: Perceived organizational support; CC: Correlation Coefficient, SEM: structural equation modelling, CB: covariance based, SPC: Standardized path coefficient, SC: Standardized coefficient

**Table 4.2.** Risk and mitigating factors for OS

| **Catego**<br>**ry** | **Factors** | **Refere**<br>**nce** | **Analysis**<br>**methods** | **Results (rounded to two decimals)** |
| --- | --- | --- | --- | --- |
| **Exacer**<br>**bating**<br>**factors** | EP and FI | Apouey<br>et al | Qualitati<br>ve | Interviews |
|  | Platform features i.e. optimization and rating | Apouey<br>et al | Qualitati<br>ve | Interviews |
|  | Urban traffic | Apouey<br>et al | Qualitati<br>ve | Interviews |
|  | MSD caused by MMH | Lin et al | T-test | The scores of job stress scale for spine disc space narrowing group are greater than normal spine group, but the differences are not significant. |
|  | Customer behaviour | Lian & Zhou | Qualitati<br>ve | Survey and in-depth interviews |
|  | High time pressure | Lian & Zhou | Qualitati<br>ve | Survey and in-depth interviews |
|  | Digitalised piece-rate | Lian & Zhou | Qualitati<br>ve | Survey and in-depth interviews |
|  | Platforms shift risks and responsibilities to couriers | Lian & Zhou | Qualitati<br>ve | Survey and in-depth interviews |
|  | Complex labour relationships | Lian & Zhou | Qualitati<br>ve | Survey and in-depth interviews |
| | Workload-Shift duration | Egozi<br>et al | Linear<br>regressio<br>n | Shift duration was significantly correlated<br>to stress ($\beta = 0.19$, $p < 0.05$). |
| | Workload-Working<br>hours | Xie,<br>Tian et<br>al | T-test/AN<br>OVA | $\leq 8$: $2.35 \pm 0.79$, $8-12$: $2.52 \pm 0.74$, $\geq 12$:<br>$2.67 \pm 0.72$ ($p < 0.01$) |
| **Amelio**<br>**rating**<br>**factors** | POS | Li et al | Multiple<br>regressio<br>n models | POS change has a significant predictive<br>effect on work performance, psychological<br>performance and behavioural performance<br>$\Delta R^2 = 0.21$ ($p < 0.001$), $0.24$ ($p < 0.001$). $0.21$<br>($p < 0.001$) |
| | EAP<br>intervention | Li et al | Multiple<br>regressio<br>n models | EAP participation can significantly lower<br>stress in three dimensions: work<br>performance, psychological performance<br>and behavioral performance, and improve<br>POS, $\Delta R^2 = 0.23$ ($p < 0.001$), $0.13$<br>($p < 0.001$), $0.21$ ($p < 0.01$) and $0.22$<br>($p < 0.001$) |
| **Socio-**<br>**demog**<br>**raphic**<br>**factors** | Gender | Xie,<br>Jiao et<br>al | T-test/AN<br>OVA | Male vs Female $2.50 \pm 0.72$ vs $2.40 \pm 0.65$<br>($p < 0.01$) |
| | | Jiang | Logistic<br>regressio<br>n | Sub-scale: PSQ, females are more likely to<br>experience personal strain (OR=1.7,<br>95%CI 1.1-2.3 $p = 0.02$) |
| | Age | Xie,<br>Jiao et<br>al | T-test/AN<br>OVA | $\leq 30$ $2.42 \pm 0.66$ vs $31-40$ $2.50 \pm 0.71$ vs $> 40$<br>$2.53 \pm 0.78$ ($p < 0.01$) |
| | Education | Jiang,<br>2017 | Logistic<br>regressio<br>n | Sub-scale PRQ: high school and above are<br>less likely to experience personal<br>resources deficiency (OR=0.4, 95%CI 0.2-0.7, $p = 0.01$) |
| | | Xie,<br>Tian et<br>al* | T-test/AN<br>OVA | Education Junior high school or below<br>$2.56 \pm 0.84$, High school $2.46 \pm 0.73$, Junior<br>college or above $2.50 \pm 0.65$ ($p < 0.05$) |
| | Marital status | Xie,<br>Tian et<br>al | T-test/AN<br>OVA | Married vs single/divorced $2.51 \pm 0.77$ vs<br>$2.45 \pm 0.70$ ($p < 0.05$) |

\* Xie, Tian et al also reported significant differences in age and gender. As its sample largely overlaps with Xie, Jiao et al, results are not included in the table.
Abbreviations: EP: Employment precarity, FI: Financial insecurity, MSD: Musculoskeletal disorder, MMH: Manual Materials Handling, POS: Perceived organizational support; EAP: Employee Assistance Program; Occupational Stress Inventory-Revised (OSI-R) has 3 sub-scales: Occupational Role Questionnaire (ORQ), Personal Strain Questionnaire (PSQ), Personal Resources Questionnaire (PRQ)

A broad range of risk and mitigating factors for couriers’ work stress were identified by quantitative and qualitative research. Papers that tested the factors using statistical methods found significant negative effects of workload such as shift duration and long daily working hours. Perceived organizational support and employer-led employee assistance program were found to have mitigating effects. Gender, age, education and marital status are the socio-demographic factors that differentiated work stress experiences. Qualitative research identified factors that could potentially contribute to OS including employment precarity, financial insecurity, customer behaviour, traffic and complex labour relationships. In addition, the use of platform technology to shift employer risks and responsibilities, intensify work and tighten managerial controls were identified. Table 4.2 summaries the relevant factors.

## DISCUSSION

This review has provided an estimate of the prevalence of couriers’ burnout and occupational stress (OS), as well as qualitatively synthesised the main risk and mitigating factors.

The six included studies on burnout reported prevalence of overall burnout or a dimension of burnout ranging from 20% to 73%. This is comparable to physicians’ burnout where overall burnout prevalence ranged from 0% to 80.5% (Rotenstein et al., 2018). Similar to this review, Rotenstein et al. (2018) did not consider the pooled results reliable due to high heterogeneity. There is considerable research on burnout among physicians, whilst couriers’ burnout is an emerging and understudied field. The six studies all used a translated or modified version of MBI to measure burnout, two of which did not report a mean score. Another two measured overall burnout whilst the last two measured one or three dimensions of burnout. These added to the heterogeneity of the results and prevented meaningful meta-analysis.

The prevalence of OS (or a dimension of OS) ranged from 7% to 90%, with a median 40%. This appeared to be relatively high compared to OS reported in other professions, for example 52.5% among health care professionals (Girma et al., 2021), 50% among medical residents (Joaquim et al., 2018), or 24.8% among university lecturers (Azizah et al., 2016). Similar to burnout, OS research in courier population is a relatively new field that needs greater attention from public health researchers. Similar to burnout research, the measurement tools used by the included studies varied significantly and the pooled result is not reliable.

Turnover intention (TI) is a psychological construct that is closely related to burnout and occupational stress (OS) and was measured in a number of studies. For example, Wen et al (Wen et al., 2020) used the Chinese version of Mobley TI scale and reported a mean score of 2.82 on a 1-5 scale. Xu et al (Xu & Zhu, 2020) used a Chinese TI scale consists of three questions, with the question -- “Looking for other job opportunities” received the highest mean score at 2.775. Xiao (Xiao, 2019) defined scores between 12 and 18 as high level of TI and the mean score was 15.27.

One included study was a longitudinal cohort study that reported an one-year intervention program (Li et al., 2021). The intervention was an Employee Assistance Program (EAP) that included a range of organizational support, such as stress management training, 7×24 counselling, forums and mini saloons, and a mental health booklet. It significantly lowered TI and OS response in the intervention group.

We found that most of the risk factors reported in the included studies are consistent with the predictors of burnout identified from broader occupational background (Meier & Kim, 2022; Shoman et al., 2021). However, as the gig economy continues to shift courier jobs to becoming digital-platform based, we found that technology issues as work stressors are not sufficiently addressed by existing literature. Both qualitative research from France (Apouey et al., 2020) and China (Lian & Zhou, 2019) found that managerial controls realised through platform technologies, such as digitalised piece rate and pay linked to customer ratings, may be associated with increased work stress among couriers. In contrast, a large number of research from sociology, human resources management, information technology and labour protection have highlighted the potential mental health risk associated with platform technology features such as gamification, algorithmic management and behavioural control (Bérastégui, 2021; Todolí-Signes, 2021; Woodcock, 2020; Woodcock & Johnson, 2018; Zhou, 2020). This factor needs to be investigated by public health research using more robust methods. It highlights an important future research area that is to examine the relationships between platform technologies and work stress.

## CONCLUSION

Based on the findings, we suggest further observational studies are needed to collect higher quality evidence. It is important to develop interventions that offer more organizational support and provide more job resources to couriers. Eight of the fifteen included studies were published in Chinese, suggesting the importance of including Chinese databases when conducting reviews in relation to couriers. We hope this review will provide a useful base for the development of research that aims to reduce work stress and burnout, and improve work-related health and safety among couriers in both developing and developed countries.

## Data Availability

All data produced in the present study are available upon reasonable request to the authors

## Acknowledgments

The authors would like to thank professor Jiangmei Qin and Dr. Yanchun Zhang of China Health Economics Association for their advices that helped refine the aims and objectives of the project.

## Contributors

MvT, HW, CA and TC conceived and designed the project. HW, SGL, TO, MKY and SL conducted reviews in English and Chinese languages and extracted the data. HW and SGL checked the data cross both languages at each steps to maintain consistent standard. SGL and SL conducted the statistical analysis to pool the prevalence. HW and TO drafted the manuscript, with SGL, CA, TC, PW and MvT contributed to the writing and editing. All remaining authors extensively reviewed and approved the final manuscript.

## Funding

The project is funded by the MRC Public Health Intervention Development Scheme (PHIND). Grant Ref: MR/T027215/1. CA is supported by NIHR Manchester Biomedical Research Centre and the NIHR Greater Manchester Patient Safety Translational Research Centre.

## Competing interests

PW is a director and shareholder of CareLoop Health Ltd, a for-profit company that develops and markets digital therapeutics for mental health conditions; and a director of Prism Life Ltd, a small research and consultancy company.

## Footnotes

1 https://www.emarketer.com/content/global-ecommerce-forecast-2021

2 https://www.emarketer.com/content/global-ecommerce-forecast-2021

3 https://www.emarketer.com/content/global-ecommerce-forecast-2021

## Notes

### Summary of Updates

Work stress has been added as another outcome of the review, in addition to burnout.

